# The Colloido-Osmotic Pressure as a useful biomarker of fatal outcome in pregnant COVID-19 patients

**DOI:** 10.1101/2023.06.29.23292050

**Authors:** B. Zavala-Barrios, A. Cérbulo-Vázquez, M. García-Espinosa, F. Caldiño-Soto, Ruth L. Madera Sandoval, L.A. Ramírez-García, I.C. Eláceo-Fernández, O. Moreno-Álvarez, G.M.L. Guerrero-Avendaño, J.C. Briones-Garduño

## Abstract

**Background:** Pregnancy was considering a health condition that could support high severity in COVID-19, among others the cardiologic and respiratory systems express some physiologic change in the pregnant women and could specially affected in COVID-19. Pregnant women characteristically have an increase volume of blood, with a higher cardiac output and lower peripheral vascular resistance, these could be negative affected by hypoalbuminemia frequently observed in COVID-19. Then, an early recognition of hypoalbuminemia in pregnant women with COVID-19 would help us to predict fatal outcome.

**Objective:** We conduct a study to analyzed the demographic, clinical, blood gas test results, laboratory and ultrasonographic doppler data in pregnant women with COVID-19 that demand medical attention for labor. Study design: Ninety-two pregnant women with COVID-19 were included in our study from may 30^th^ 2020 to august 12^th^ 2022, and data was analyzed in survival and fatal outcome patients. Normality test were applied to data and parametric or non-parametric test were used to determine statistical difference between or among data.

**Results:** Demographic and clinical data were quite similar between survival and fatal outcome pregnant patients with COVID-19. Also, blood gas test shown similar results among groups (asymptomatic, mild, or severe patients who survive COVID-19 or fatal outcome patients). We observed that serum albumin was consistently low in pregnant patients with fatal outcome by COVID-19, then based on Total Protein concentration in serum we calculated the colloido-osmotic pressure and we evaluate its utility as predictor of fatal outcome. ROC analysis and Kaplan-Meier of survival curves suggest that the colloido-osmotic pressure could be a potential predictor. Also, we observed an association of low colloido-osmotic pressure and low amniotic fluid index.

**Conclusion:** Our study suggests that early evaluation of pregnant with COVID-19 must include the calculation of colloido-osmotic pressure and the doppler analysis to early recognition of fatal outcome in pregnant women with COVID-19.

## Introduction

Since 2019 the high number of persons with SARS-CoV-2 result in a pandemic emergency with loss of millions of lives ^1^. Fatal outcome by COVID-19 has been associated with certain comorbidities such as obesity or hypertension among others, also pregnant women may experience a high fatality rate probably because a high susceptibility to inflammation ^2^. During pregnancy, the immune response develops an immunological tolerance and a high level of regulation to inflammatory response, however this delicate balance could negative affected in pregnant women with COVID-19 ^3^. Also, a profound physiologic regulation in other systems can be observed in pregnancy, which includes the circulatory and respiratory systems ^4,5^. Pregnant women characteristically have an increase volume of blood, with a higher cardiac output and lower peripheral vascular resistance, also, pregnant women tend to develop a respiratory alkalosis with pH between normal limits, and all these characteristics could be negative affected by COVID-19.

COVID-19-associated deaths during pregnancy was increasing worldwide, Karimi et al reported 153 deaths out of 11758 pregnant and postpartum women with COVID-19 (1.3%). However, the mortality rate is differential between high-or middle-income countries. The high-income countries show a mortality rate about 0.19%, while in middle-income countries was as high as 8.51% ^6^. In Mexico, from April 2022 to August 2021, 389 maternal deaths by COVID-19 were recorded in a population of 20329 pregnant and postpartum women (1.9%) ^7^.

Demographic data in pregnant women with COVID-19 shows that they are frequently older than 31 years old ^8^, with a Body Mass Index >30, multiple pregnancy and hypertension ^8-10^. In addition, some social condition in mild-income countries could lead to undernutrition especially in pregnant women, and leading to poor immune response against COVID-19 ^11,12^. Undernutrition could be recognized by low serum albumin, which is a good predictor for negative outcomes after surgeries ^13^, or infection disease as COVID-19 ^14^.

In addition, certain laboratory parameters in pregnant women with COVID-19 are useful to detect high risk of fatal outcome, among these parameters the leucocyte count ^15^, platelet count ^16^ or neutrophil-lymphocyte ratio (NLR)^17^ had been reported. Likewise, the evaluation of arterial blood gas parameters is essential to evaluate pregnant women with COVID-19, however, a low number of studies shows the arterial blood gas test in pregnant women with COVID-19. Also, it has been reported that some chemistry parameters can be useful for early detection of severe COVID-19, among them the serum values of creatinine ^18^, procalcitonin (PCT) ^19^ or albumin ^20^ had been reported, however, poor analysis is available for pregnant women with COVID-19. Specially the hypoalbuminemia could result in negative changes in pregnant women because it led to a low Colloido-Osmotic Pressure (cCOP). This is a pressure exerted by the protein on the capillary membranes, and could be calculated using the formula 1 of Landis-Pappenheimer, and fatal outcome in trauma patients is frequently observed in patients with values as low as <15 mmHg of cCOP ^21^. A low cCOP could limit the maternal-fetal interchange, compromise the placental perfusion and altering the microvasculature in chorionic villi ^22^, however, all these changes could closely monitored by doppler ultrasonography, following the blood flow in the: Umbilical artery (UA), uterine artery (UAT), Fetal Middle Cerebral Artery (FMCA), or Amniotic Fluid Index (AFI). Until now, there are few studies that include the ultrasonographic evaluation in pregnant women with COVID-19, and the few reported no differences between SARS-CoV-2-positive pregnant women and controls ^23^.

Our study analyzed demographic, clinical, laboratory, arterial blood gas test and doppler data in pregnant women with COVID-19 who survive or got a fatal outcome, the results suggest that cCOP can early detect pregnant women with COVID-19 with high probability of having a fatal outcome.

## Material and methods

### Patients

This study was conducted by the “Servicio de Ginecología y Obstetricia” at the Hospital General de Mexico “Dr. Eduardo Liceaga” (HGM, Research project: *DI/20112/04/45*), and the Servicio de complicaciones de la segunda mitad del embarazo, División Obstetricia, UMAE Hospital de Gineco-Obstetricia No. 4 “Dr. Luis Castelazo Ayala” (UMAE HGO-4 IMSS. Research project: *R-2020-785-095*). After obtaining a signed informed consent letter, ninety-two women were enrolled and assigned to one of three groups: a) asymptomatic, n=20, b) mild COVID-19, n=56, c) severe COVID-19, n=16, then 6 patients in the severe COVID-19 group got a fatal outcome and was analyzed as a different group. The COVID-19 diagnosis was based on clinical characteristics and positive RT-PCR test for SARS-CoV-2 or radiographic images compatible with COVID-19 disease. Demographic, Clinical and Comorbidities features were recorded.

### Blood sample collection

Our study is in accordance with the World Medical Association’s Declaration of Helsinki. After the patients signed the informed consent letter, blood specimens were collected in silicone-coated tubes (EDTA or heparinized tubes, BD Vacutainer, N. J, USA), samples were processed immediately after collection.

### Arterial Blood Gas test

Heparin blood samples were obtained and blood gas analysis was performed on an ABL90 Flex Plus blood gas analyzer (Radiometer Medical ApS, Bronshoj, Denmark) within 30 min of collect.

### Doppler test

Doppler analysis was as DÁntonio reported ^24^, we performed the analysis using a Samsung HS40 (Samsung Healthcare, N.J, USA) or a Chison EBit 50 (Chison, Jiangsu, China) ultrasound machines. Data was recorded during fetal quiescence, without fetal tachycardia, with the insonation angle kept as close to 0° as possible. FMCA was examined at the point at which it passes the sphenoid wing, close to the circle of Willis, and the UA was examined at a free loop of the umbilical cord. To record the UAT Doppler, the transducer was placed over the iliac fossa and the course of the uterine arteries was followed from the lateral pelvic wall across external iliac artery using color Doppler. Pulsed Doppler was then applied 1 cm medial to the crossover point. When three similar consecutive waveforms were obtained, the PI was measured, and the mean value of the left and right uterine artery PI calculated.

### Statistical analysis

Statistical analysis was performed using GraphPad Prism® version 7 software (GraphPad Software, San Diego, CA, USA) and the R i386 3.5.2 terminal (Microsoft corp., Boston, MA, USA.). Categorical variables were expressed as percentages (%), and compared by Fisher’s exact test. Non-parametric ANOVA (Kruskal–Wallis test) with Dunn’s posttest was applied. Spearman correlation for interest variables was implemented. A *p*<0.05 was considered statistically significant.

## Results

Ninety-two pregnant patients with COVID-19 was attended in the HGM and reviewed with coauthors in the UMAE HGO-4 IMSS, from may 30^th^ 2020 to august 12^th^ 2022. All patients got an RT-PCR+ for SARS-CoV-2 or radiographic imagen diagnostic for SARS-CoV-2 infection. Twenty of the ninety-two pregnant patients in labor without any clinical sign of COVID-19 got a positive result for SARS-CoV-2 PCR test, these asymptomatic patients were included in our study. Table 1 show clinical and laboratory characteristics, no statistical difference was observed for age, body mass index, gestational age, trimester of gestation, weight newborn, or Apgar score among the groups, however, prolonged stays in hospital were observed in severe or fatal outcome patients with COVID-19 than in asymptomatic patients positive for SARS-CoV-2. Also, we observed a similar number of leucocytes and platelets among groups (Table 1). However, the percentage of PMN was higher in patients with severe COVID-19 than in asymptomatic pregnant women.

**Table 1.**
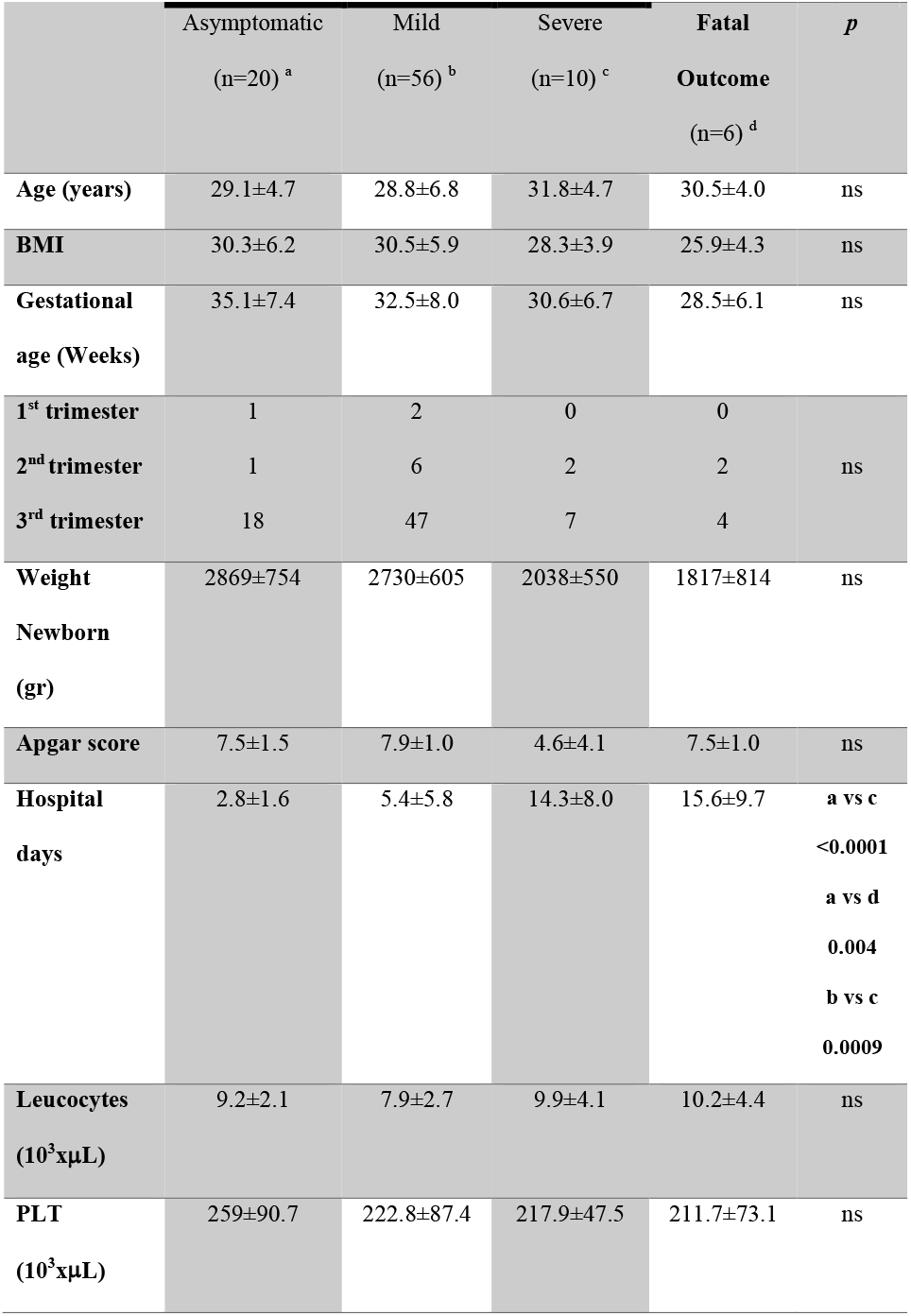
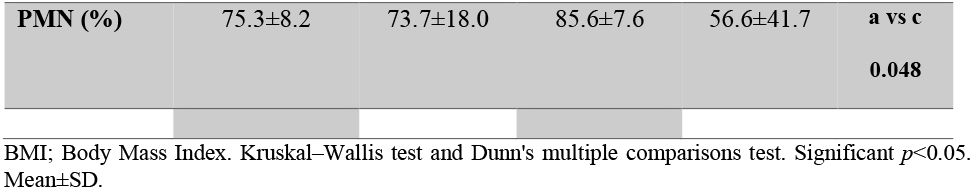
Clinical and laboratory characteristics of pregnant women with COVID-19.

Table 2 shows the results of arterial blood gas test. No statistical significance was observed in pH, PaO_2_, saturation of O_2_ or lactate among groups, however, severe patients show a higher CO_2_ value than mild patients (p=0.005), also, the BE was lower in the mild group than in fatal outcome patients (p=0.02). Results in Table 2 shows that pregnant women developing a respiratory alkalosis probably due by hyperventilation, this respiratory condition is quite frequent in pregnant patients and pH values are in normal limits.

**Table 2.**
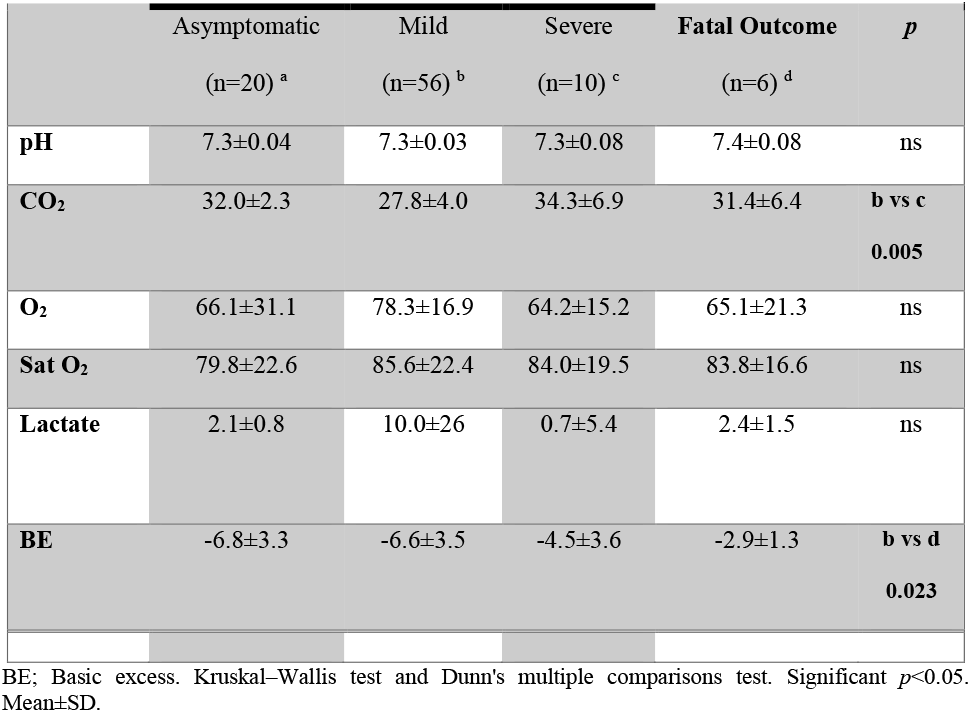
Arterial Blood Gas parameters of pregnant women with COVID-19.

Regarding chemistry parameters, we observed that creatinine and D-Dimer concentration were similar among groups (Table 3), however, procalcitonin was higher in severe COVID-19 than in asymptomatic patients (p=0.040). Also, the asymptomatic patients show the highest serum albumin concentration, while the lowest serum albumin was in fatal outcome pregnant women with COVID-19 (Table 3).

**Table 3.** Clinical chemistry parameters of pregnant women with COVID-19.

|  | Survivor |  |  |  | *p* |
| --- | --- | --- | --- | --- | --- |
|  | Asymptomatic<br>(n=20) <sup>a</sup> | Mild<br>(n=56) <sup>b</sup> | Severe<br>(n=10) <sup>c</sup> | Fatal Outcome<br>(n=6) <sup>d</sup> |  |
| **Creatinine**<br>**(mg/dL)** | 0.5±0.08 | 0.2±1.7 | 0.8±0.8 | 0.5±0.2 | ns |
| **D-Dimer**<br>**(ng/mL)** | 2261±1254 | 2022±2035 | 3303±3825 | 11360±17192 | ns |
| PCT<br>(ng/mL) | 0.07±0.06 | 496.2±1129 | 1.3±2.7 | 0.3±0.1 | a vs c<br>0.040 |
| Albumin<br>(g/dL) | 3.2±0.2 | 3.1±0.4 | 2.7±0.3 | 2.6±0.1 | a vs c<br>0.021<br>a vs d<br>0.005<br>b vs c<br>0.036<br>b vs d<br>0.010 |

PCT; Procalcitonin. Kruskal–Wallis test and Dunn's multiple comparisons test. Significant $p<0.05$ . Mean±SD.

Fig. 1A shows the cCOP in mild-severe COVID-19 pregnant women is higher than in fatal outcome pregnant women with COVID-19. Moreover, moderate correlation (r= 0.45) among cCOP and amniotic fluid index, was observed (fig.1B) in patients with mild, severe and fatal outcome.

**Fig. 1.**
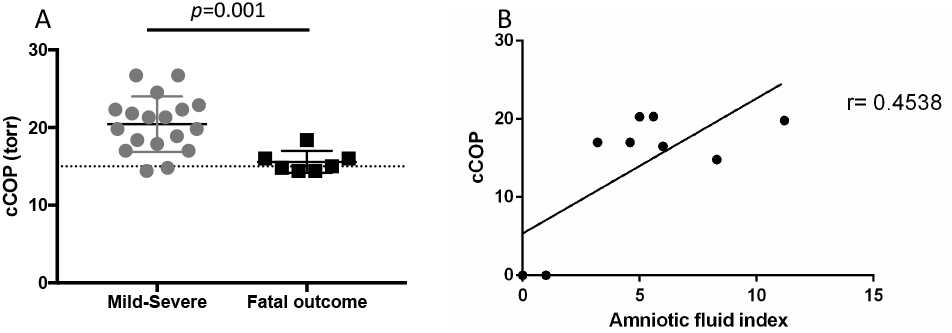
Colloido-Osmotic Pressure calculated in severe and fatal outcome pregnant women with COVID-19. The cCOP was calculated as described in the methods. A) Pregnant women with Mild-Severe COVID-19 have a higher cCOP than in pregnant women with fatal outcome, B) Sperman correlation between cCOP and amniotic fluid index. The correlation value (among mild, severe COVID-19 and fatal outcome patients) suggesting a medium correlation, p= 0.22. The results are expressed as the mean±SD. Significance value was *p*<0.05. U Mann-Whitney tests were calculated.

Also, the ROC curve analysis show that the AUC was 53.2%, and the optimal cutoff was calculated in accordance with Youden index 15.7, with a confidence interval of 77.8%, 57.1%. Also, a survival curve of cCOP was analyzed. Kaplan-Meier of survival curves were made according to the optimal cutoff, and hazard ratio expressed in percentage, *p*<0.05. The median days of survival was 340 for the lower risk group and a 31% of risk for the patients with value greater than 15.7.

Fig. 2 shows the ultrasonographic analysis in survivor pregnant women with COVID-19 or asymptomatic patients SARS-CoV-2 positive. Similar results were observed for UA, UAT, and FMCA among groups, however, the amniotic fluid index show statistical difference, were asymptomatic patients show a higher index than in severe COVID-19 patients (p=0.01) or higher index in mild COVID-19 patients than in severe COVID-19 patients (p=0.02).

**Fig. 2.**
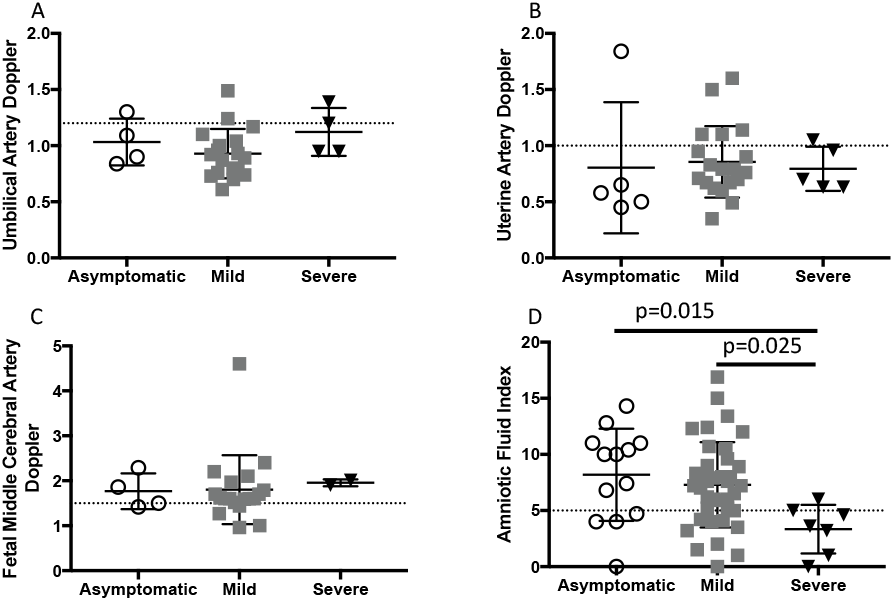
Maternal-fetal Doppler test in pregnant women with COVID-19. Pregnant women were tested by USG as described in the methods. Doppler analysis is shown for A) Umbilical artery, B) Uterine artery, C) Fetal middle cerebral artery, and D) Amniotic fluid index. Dash line indicate normal limits. The results are expressed as the mean±SD. Significance value was *p*<0.05. Kruskal–Wallis and Dunn’s multiple comparisons tests were calculated.

## Discussion

### Principal Findings

The early recognition of severe or critical COVID-19 in pregnant women would help us to get a lower maternal mortality by COVID-19, or predict fatal outcome in pregnant women with COVID-19. Our study shows that long stay in hospital (media=14-15 days) is characteristic of patients with severe COVID-19 or fatal outcome (Table 1). We also observed that arterial blood gas test was quite similar among pregnant women with COVID-19 even in those who got a fatal outcome (Table 2). In agreement with the literature hypoalbuminemia was noted specially in patients with severe COVID-19 or those who got fatal outcome (Table 3). We calculated the Colloido-osmotic pressure in mild and severe COVID-19 and compared with patients with fatal outcome, a significant difference was observed between those groups, and association between cCOP and amniotic fluid index was observed (Figure 1). Unfortunately, a doppler test was not applied on patients with fatal outcome by COVID-19, however, we observed that the amniotic fluid index was lower according to severity of COVID-19 (Figure 2).

### Results

Several studies show the mean of age of pregnant women with COVID-19 is around 30 years old ^8,15,25-31^, and most pregnant women with COVID-19 were aged between 20 to 39 years ^9^, our study also show that pregnant women with COVID-19 or asymptomatic patients were around 30 years old (Table 1). Comorbidities as obesity has been associated to suffer severe COVID-19 in pregnant women ^8^, and been in a higher risk for maternal death ^32^, and some studies show that BMI over 30 is frequently observed in pregnant women COVID-19 patients ^28,31^, our study also observed this condition (Table 1). The most of literature show that pregnant women with COVID-19 have a mean of gestational age around 30 weeks ^28,29^, or higher gestational age than 30 weeks ^26,27^, as we expect the most of maternal death in pregnant women with COVID-19 has been observed in the third trimester of pregnancy, however, other studies did not show a high number of fatal outcomes by COVID-19 in the third trimester ^33-36^. Maternal mortality could be as low as 0.2 to 2% ^10,37-39^, or as high as 10 to 20% ^6,40-42^ according with low-or middle-income ^42-44^. Our study shows similar data for gestational age, fatal outcome by trimester and maternal mortality as a middle-income country, these observation in a sense validate our study and could help to increase the number of observations for pregnant women with COVID-19 in the very first wave when collection of data was so difficult.

Early evaluation of blood gas test in pregnant women with COVID-19 seems to be critical and could help to recognize severity. Both alkalosis and acidosis were observed in patients with COVID-19 in general population, and alkalosis was observed in 54.3% of the subjects ^45^. Also, ketoacidosis has been reported in pregnant women with COVID-19 which is associated with diabetes, hyperglycemia and ketosis ^46,47^. However, our result did not show great differences among groups, indicating that blood gas parameters was not altered even with variants of SARS-CoV-2 that express high maternal mortality. More observations must do it to clarify this observation.

Blood cell count had been analyzed in patients with COVID-19, some authors show that white blood cell count is higher in pregnant than in non-pregnant women with COVID-19 ^27^, however, other studies show that leucocytosis was not observed in pregnant women with COVID-19 ^27,35^, in contrast, some pregnant women with COVID-19 show lymphopenia ^48^. In agreement with Wang Z et. al. and Yan J et. al. we did not observe great differences in leucocytes counts among the groups. Finally, has been reported that a lower platelet count is observed in patients with severe COVID-19 ^16,18^, while other study show that the platelet count was similar in pregnant women with and without SARS-CoV-2 infection ^49^. In agreement with Covali R et. al. we did not observed difference in the platelet count of pregnant women with COVID-19, no difference was observed among groups by severity or fatal outcome, this is probably because the relatively small number of patients. More studies are necessary to clarify if low platelet count is a good predictor of fatal outcome in pregnant women.

Shmakov RG et. al. shows a similar D-Dimer concentration among asymptomatic, mild, moderate and severe COVID-19 in pregnant women ^28^, and our study agrees with this observation. Another potential biomarker is the PCT, however, the value of this biomarker to predict severity in COVID-19 show some contradictory results in general population ^19^, and in pregnant women with COVI-19, PCT levels was similar between pregnant and non-pregnant women with COVID-19 ^41,50^, while some other study show a difference between them ^27^. Because the PCT was similar among mild, moderate and severe COVID-19 in pregnant women ^28^, the result suggest that PCT is not able to discriminate the mild from severe COVID-19 in pregnant women, our study agrees with this observation, we observed that the highest PCT concentration was in the mild cases, in the mild group we observed patients with PCT values quite similar to severe COVID-19 patients (range 0.010-11.0 ng/mL) and a subgroup of mild COVID-19 patients with high values of PCT (918.0-5227.0 ng/mL), suggesting that this second subgroup got a bacterial infection, however, we could not get a positive bacterial culture on them, interestingly, none of this potential over infected patients got a fatal outcome, suggesting that even in the case of double infection the immune system of pregnant women will be sufficient to solve the infection challenge.

Hypoalbuminemia has been associated to disease severity in COVID-19 ^20,28,51^, the expression of hypoalbuminemia on admission is more frequent in non-survivors than survivors, and could be useful as an early predictor of mortality in COVID-19 ^52-54^. In contrast, a high level of albumin on admission is associated with a lower risk of Acute Respiratory Distress Syndrome and admission to ICU ^53^. Lower albumin concentration was reported in pregnant than in non-pregnant women with COVID-19 without fatal outcome ^27^, our results show that a hypoalbuminemia as low as 2.5 could result in low colloido-osmotic pressure and finally a fatal outcome. The colloido-osmotic pressure is easily evaluated among patients, and our study propose its routine analysis to predict fatal outcome in pregnant women with COVID-19. Moreover, low cCOP in COVID-19 could lead to impaired placental perfusion. It has been reported that SARS-CoV-2 infection does not affect the blood flow in the umbilical vein ^55^, also, in pregnant women recovered from COVID-19 the pulsatility and resistance indices of umbilical and uterine artery show a significant increase than in controls ^56^. In contrast, Soto-Torres et. al. report that Doppler evaluation was similar between pregnant women with SARS-CoV-2 infection and controls ^23^. In agreement with Soto-Torres et. al. we observed that the umbilical artery, uterine artery, and fetal middle cerebral artery the blood flow were similar in pregnant and non-pregnant COVID-19 women, however, we observed that the Amniotic Fluid Index was lower in severe than in mild or asymptomatic COVID-19 patients, this lower amniotic fluid index in severe COVID-19 patients could expressed or been the Fetal Inflammatory Response Syndrome by viral infection.

### Clinical implications

Two conditions could explain the high maternal mortality in our hospital a) because we are in a middle-income country and b) because our hospital is a national reference center. We were able to classify our patients in asymptomatic, mild, and severe COVID-19 patients; however, some clinical characteristics of our patients were quite similar among groups as age, BMI, gestational age, weight newborn, Apgar score etc. indicating that those characteristics have low value to early recognize patients with high potential for a fatal outcome. Also, our study show that pregnant women tend to respiratory alkalosis that is not necessarily deteriorated by COVID-19. Despite a limited respiratory condition in pregnant women, it seems that gas test in pregnant women with COVID-19 is similar than in women in general population with COVID-19. Some other laboratory analysis as the blood count was also analyzed, we did not observe leukocytosis or lymphopenia in pregnant women with COVID-19, and no association with severity was observed, probably because a small number of patients in our study. We observed the highest neutrophilia in pregnant women with severe COVID-19, however, neutropenia and not neutrophilia was observed in pregnant women with COVID-19 and fatal outcome. Neutrophils are important innate cells that participate in many mechanisms related with inflammation, its high or low number in pregnant women with COVID-19 seems to be important for limit the infection, however, we need more analysis in pregnant women to clarify if neutropenia and neutrophilia are supporter of pathophysiology in COVID-19. Our results suggest that Basic Doppler evaluation could be useful to early recognition of severe COVID-19 in pregnant women.

### Research implications

We do not know when the next pandemic comes, but patients with comorbidities or special conditions as pregnancy will be affected. Due the complex condition in prenatal surveillance, labor and postpartum period during pandemic for COVID-19 many opportunities of do deeper analysis of data o searching for a no-conventional assessment were lost, and data in pregnant women with COVID-19 is scarce. Medical attention of pregnant patients must include a holistic view, with strict attention to ethical and patient’s rights, and then collect any data that potentially could help to understand the new disease like COVID-19 and minimize the fatal outcomes.

### Strengths and limitations

Our study has some limitations and strengths, the number of observations is relatively low (n=92) with six fatal outcomes included (6.5%), because early in the pandemic a medical treatment was not established and medical support was apply after some days of the initial COVID-19 symptoms a high percentage of fatal outcome was observed, however, our study also show demographic, clinical and laboratory data observed at the very early time (in the second wave) of pandemic of COVID-19 in Mexico, and when vaccination was not available, these conditions could help us to understand the natural progression in pregnant women with COVID-19.

### Conclusions

A high number of comorbidities is observed in COVID-19 patients with high severity, and also the pregnancy could allow a severe presentation of COVID-19. However, the frequency of severe cases in pregnant women with COVID-19 is similar than in general population ^15,41,57^, due to the pandemic condition many studies in pregnant women with COVID-19 show missing or not collected data, our data show that hypoalbuminemia and the cCOP could help to early recognize patients with high risk of fatal outcome, more studies are necessary to know if pregnancy must not take as a comorbidity that increase the risk of severe COVID-19.

## Data Availability

All data produced in the present study are available upon reasonable request to the authors

## Contributors

BZB, ACV and JCBG conceived and designed the study and contributed to data analysis. BZB and ACV wrote the first version of manuscript. MGE, FCS, LARG, RLMS, JCBG, OMA, and GMLGA contributed to a critical revision of the report. MGE, FCS, LARG, ICEF, JCBG, BZB, and OMA contributed to the clinical evaluation of patients and supervision of medical treatments and patient care. RLMS contributed to do the statistical analysis and graph data. All authors reviewed the final version. JCBG reviewed and approved the final version.

## Acknowledgments

The authors extend gratitude to the staff at Gynecology & Obstetrics Department in the General Hospital of Mexico “Dr. Eduardo Liceaga” and Gynecology & Obstetric Hospital No. 4 UMAE “Dr. Luis Castelazo Ayala”.

## References

1. WHO. WHO Coronavirus (COVID-19) Dashboard. https://covid19.who.int/

2. Adab P, Haroon S, O’Hara ME, Jordan RE. Comorbidities and covid-19. BMJ. Jun 15 2022;377:o1431. doi:10.1136/bmj.o1431

3. Cerbulo-Vazquez A, Garcia-Espinosa M, Briones-Garduno JC, et al. The percentage of CD39+ monocytes is higher in pregnant COVID-19+ patients than in nonpregnant COVID-19+ patients. PLoS One. 2022;17(7):e0264566. doi:10.1371/journal.pone.0264566

4. Hegewald MJ, Crapo RO. Respiratory physiology in pregnancy. Clin Chest Med. Mar 2011;32(1):1–13. doi:10.1016/j.ccm.2010.11.001

5. LoMauro A, Aliverti A. Respiratory physiology in pregnancy and assessment of pulmonary function. Best Pract Res Clin Obstet Gynaecol. Jun 27 2022;doi:10.1016/j.bpobgyn.2022.05.007

6. Karimi L, Makvandi S, Vahedian-Azimi A, Sathyapalan T, Sahebkar A. Effect of COVID-19 on Mortality of Pregnant and Postpartum Women: A Systematic Review and Meta-Analysis. J Pregnancy. 2021;2021:8870129. doi:10.1155/2021/8870129

7. Epidemiología CNdEdGySRDGd. Aviso Epidemiologico COVID-19. https://www.gob.mx x› uploads › attachment › file

8. Epelboin S, Labrosse J, De Mouzon J, et al. Obstetrical outcomes and maternal morbidities associated with COVID-19 in pregnant women in France: A national retrospective cohort study. PLoS Med. Nov 2021;18(11):e1003857. doi:10.1371/journal.pmed.1003857

9. Galang RR, Newton SM, Woodworth KR, et al. Risk Factors for Illness Severity Among Pregnant Women With Confirmed Severe Acute Respiratory Syndrome Coronavirus 2 Infection-Surveillance for Emerging Threats to Mothers and Babies Network, 22 State, Local, and Territorial Health Departments, 29 March 2020-5 March 2021. Clin Infect Dis. Jul 15 2021;73(Suppl 1):S17–S23. doi:10.1093/cid/ciab432

10. Overton EE, Goffman D, Friedman AM. The Epidemiology of COVID-19 in Pregnancy. Clin Obstet Gynecol. Mar 1 2022;65(1):110–122. doi:10.1097/GRF.0000000000000674

11. James PT, Ali Z, Armitage AE, et al. The Role of Nutrition in COVID-19 Susceptibility and Severity of Disease: A Systematic Review. J Nutr. Jul 1 2021;151(7):1854–1878. doi:10.1093/jn/nxab059

12. Don BR, Kaysen G. Serum albumin: relationship to inflammation and nutrition. Semin Dial. Nov-Dec 2004;17(6):432–7. doi:10.1111/j.0894-0959.2004.17603.x

13. Gibbs J, Cull W, Henderson W, Daley J, Hur K, Khuri SF. Preoperative serum albumin level as a predictor of operative mortality and morbidity: results from the National VA Surgical Risk Study. Arch Surg. Jan 1999;134(1):36–42. doi:10.1001/archsurg.134.1.36

14. Mentella MC, Scaldaferri F, Gasbarrini A, Miggiano GAD. The Role of Nutrition in the COVID-19 Pandemic. Nutrients. Mar 27 2021;13(4) doi:10.3390/nu13041093

15. Amini Moghadam S, Dini P, Nassiri S, Motavaselian M, Hajibaba M, Sohrabi M. Clinical features of pregnant women in Iran who died due to COVID-19. Int J Gynaecol Obstet. Feb 2021;152(2):215–219. doi:10.1002/ijgo.13461

16. Lippi G, Plebani M, Henry BM. Thrombocytopenia is associated with severe coronavirus disease 2019 (COVID-19) infections: A meta-analysis. Clin Chim Acta. Jul 2020;506:145–148. doi:10.1016/j.cca.2020.03.022

17. Lasser DM, Chervenak J, Moore RM, et al. Severity of COVID-19 Respiratory Complications during Pregnancy are Associated with Degree of Lymphopenia and Neutrophil to Lymphocyte Ratio on Presentation: A Multicenter Cohort Study. Am J Perinatol. Oct 2021;38(12):1236–1243. doi:10.1055/s-0041-1732421

18. Tufa A, Gebremariam TH, Manyazewal T, et al. Limited value of neutrophil-to-lymphocyte ratio and serum creatinine as point-of-care biomarkers of disease severity and infection mortality in patients hospitalized with COVID-19. PLoS One. 2022;17(10):e0275391. doi:10.1371/journal.pone.0275391

19. Wolfisberg S, Gregoriano C, Schuetz P. Procalcitonin for individualizing antibiotic treatment: an update with a focus on COVID-19. Crit Rev Clin Lab Sci. Jan 2022;59(1):54–65. doi:10.1080/10408363.2021.1975637

20. Johnson AS, Winlow W. COVID-19 vulnerabilities are intensified by declining human serum albumin levels. Exp Physiol. Jul 2022;107(7):674–682. doi:10.1113/EP089703

21. Alberto Basilio Olivares JCB, Jesús Antonio Jiménez Origel,, Ponce MADdL. La presión coloidosmótica (PCO) como indicador pronóstico en trauma. Reporte preliminar. Rev Asoc Mex Med Crit y Ter Int. 2012;26(4):230–233.

22. Acharya G, Sonesson SE, Flo K, Rasanen J, Odibo A. Hemodynamic aspects of normal human feto-placental (umbilical) circulation. Acta Obstet Gynecol Scand. Jun 2016;95(6):672–82. doi:10.1111/aogs.12919

23. Soto-Torres E, Hernandez-Andrade E, Huntley E, Mendez-Figueroa H, Blackwell SC. Ultrasound and Doppler findings in pregnant women with SARS-CoV-2 infection. Ultrasound Obstet Gynecol. Jul 2021;58(1):111–120. doi:10.1002/uog.23642

24. D’Antonio F, Rizzo G, Gustapane S, et al. Diagnostic accuracy of Doppler ultrasound in predicting perinatal outcome in pregnancies at term: A prospective longitudinal study. Acta Obstet Gynecol Scand. Jan 2020;99(1):42–47. doi:10.1111/aogs.13705

25. Dawood FS, Varner M, Tita A, et al. Incidence and Clinical Characteristics of and Risk Factors for Severe Acute Respiratory Syndrome Coronavirus 2 (SARS-CoV-2) Infection Among Pregnant Individuals in the United States. Clin Infect Dis. Jul 6 2022;74(12):2218–2226. doi:10.1093/cid/ciab713

26. Zhang J, Zhang Y, Ma Y, et al. The associated factors of cesarean section during COVID-19 pandemic: a cross-sectional study in nine cities of China. Environ Health Prev Med. Oct 10 2020;25(1):60. doi:10.1186/s12199-020-00899-w

27. Wang Z, Wang Z, Xiong G. Clinical characteristics and laboratory results of pregnant women with COVID-19 in Wuhan, China. Int J Gynaecol Obstet. Sep 2020;150(3):312–317. doi:10.1002/ijgo.13265

28. Shmakov RG, Prikhodko A, Polushkina E, et al. Clinical course of novel COVID-19 infection in pregnant women. J Matern Fetal Neonatal Med. Dec 2022;35(23):4431–4437. doi:10.1080/14767058.2020.1850683

29. Pierce-Williams RAM, Burd J, Felder L, et al. Clinical course of severe and critical coronavirus disease 2019 in hospitalized pregnancies: a United States cohort study. Am J Obstet Gynecol MFM. Aug 2020;2(3):100134. doi:10.1016/j.ajogmf.2020.100134

30. Capobianco G, Saderi L, Aliberti S, et al. COVID-19 in pregnant women: A systematic review and meta-analysis. Eur J Obstet Gynecol Reprod Biol. Sep 2020;252:543–558. doi:10.1016/j.ejogrb.2020.07.006

31. Afshar Y, Gaw SL, Flaherman VJ, et al. Clinical Presentation of Coronavirus Disease 2019 (COVID-19) in Pregnant and Recently Pregnant People. Obstet Gynecol. Dec 2020;136(6):1117–1125. doi:10.1097/AOG.0000000000004178

32. Mendez-Dominguez N, Santos-Zaldivar K, Gomez-Carro S, Datta-Banik S, Carrillo G. Maternal mortality during the COVID-19 pandemic in Mexico: a preliminary analysis during the first year. BMC Public Health. Jul 2 2021;21(1):1297. doi:10.1186/s12889-021-11325-3

33. Gupta V, Yadav Y, Sharma R, Mishra M, Ambedkar D, Gupta V. Maternal and Perinatal Outcomes of Hospitalized COVID-19 Positive Pregnant Women. Cureus. Feb 2022;14(2):e21817. doi:10.7759/cureus.21817

34. DeBolt CA, Bianco A, Limaye MA, et al. Pregnant women with severe or critical coronavirus disease 2019 have increased composite morbidity compared with nonpregnant matched controls. Am J Obstet Gynecol. May 2021;224(5):510 e1–510 e12. doi:10.1016/j.ajog.2020.11.022

35. Yan J, Guo J, Fan C, et al. Coronavirus disease 2019 in pregnant women: a report based on 116 cases. Am J Obstet Gynecol. Jul 2020;223(1):111 e1–111 e14. doi:10.1016/j.ajog.2020.04.014

36. London V, McLaren R, Jr., Atallah F, et al. The Relationship between Status at Presentation and Outcomes among Pregnant Women with COVID-19. Am J Perinatol. Aug 2020;37(10):991–994. doi:10.1055/s-0040-1712164

37. Metz TD, Clifton RG, Hughes BL, et al. Association of SARS-CoV-2 Infection With Serious Maternal Morbidity and Mortality From Obstetric Complications. JAMA. Feb 22 2022;327(8):748–759. doi:10.1001/jama.2022.1190

38. Borges Charepe N, Queiros A, Alves MJ, et al. One Year of COVID-19 in Pregnancy: A National Wide Collaborative Study. Acta Med Port. May 2 2022;35(5):357–366. doi:10.20344/amp.16574

39. Juan J, Gil MM, Rong Z, Zhang Y, Yang H, Poon LC. Effect of coronavirus disease 2019 (COVID-19) on maternal, perinatal and neonatal outcome: systematic review. Ultrasound Obstet Gynecol. Jul 2020;56(1):15–27. doi:10.1002/uog.22088

40. Leung C, Simoes ESAC, Oliveira EA. Are in-hospital COVID-19-related mortality and morbidity in pregnancy associated with gestational age? Ultrasound Obstet Gynecol. Aug 2022;60(2):234–242. doi:10.1002/uog.24931

41. Khoiwal K, Ravi AK, Arora S, et al. Impact of Pregnancy on Susceptibility and Severity of COVID-19: A Hospital-Based Prospective Observational Study. Cureus. Apr 2022;14(4):e24281. doi:10.7759/cureus.24281

42. Nakamura-Pereira M, Betina Andreucci C, de Oliveira Menezes M, Knobel R, Takemoto MLS. Worldwide maternal deaths due to COVID-19: A brief review. Int J Gynaecol Obstet. Oct 2020;151(1):148–150. doi:10.1002/ijgo.13328

43. Takemoto MLS, Menezes MO, Andreucci CB, et al. Maternal mortality and COVID-19. J Matern Fetal Neonatal Med. Jun 2022;35(12):2355–2361. doi:10.1080/14767058.2020.1786056

44. Maza-Arnedo F, Paternina-Caicedo A, Sosa CG, et al. Maternal mortality linked to COVID-19 in Latin America: Results from a multi-country collaborative database of 447 deaths. Lancet Reg Health Am. Aug 2022;12:100269. doi:10.1016/j.lana.2022.100269

45. Sanghani H, Bansal S, Parmar V, Shah R. Study of Arterial Blood Gas Analysis in Moderate-to-Severe COVID-19 Patients. Cureus. Jul 2022;14(7):e26715. doi:10.7759/cureus.26715

46. van Amesfoort JE, Werter DE, Painter RC, Hermans FJR. Severe metabolic ketoacidosis as a primary manifestation of SARS-CoV-2 infection in non-diabetic pregnancy. BMJ Case Rep. Apr 19 2021;14(4) doi:10.1136/bcr-2021-241745

47. Pikovsky M, Tan MY, Ahmed A, Sykes L, Agha-Jaffar R, Yu CKH. Euglycaemic ketoacidosis in pregnant women with COVID-19: two case reports. BMC Pregnancy Childbirth. Jun 16 2021;21(1):427. doi:10.1186/s12884-021-03928-w

48. Al-Saadi E, Abdulnabi MA. Hematological changes associated with COVID-19 infection. J Clin Lab Anal. Jan 2022;36(1):e24064. doi:10.1002/jcla.24064

49. Covali R, Socolov D, Socolov R, et al. Complete Blood Count Peculiarities in Pregnant SARS-CoV-2-Infected Patients at Term: A Cohort Study. Diagnostics (Basel). Dec 30 2021;12(1) doi:10.3390/diagnostics12010080

50. Asghar MS, Siddiqui MA, Iqbal S, et al. COVID-19 infection among pregnant and non-pregnant women: Comparison of biochemical markers and outcomes during COVID-19 pandemic, A retrospective cohort study. Ann Med Surg (Lond). Apr 2022;76:103527. doi:10.1016/j.amsu.2022.103527

51. Xu Y, Yang H, Wang J, et al. Serum Albumin Levels are a Predictor of COVID-19 Patient Prognosis: Evidence from a Single Cohort in Chongqing, China. Int J Gen Med. 2021;14:2785–2797. doi:10.2147/IJGM.S312521

52. Viana-Llamas MC, Arroyo-Espliguero R, Silva-Obregon JA, et al. Hypoalbuminemia on admission in COVID-19 infection: An early predictor of mortality and adverse events. A retrospective observational study. Med Clin (Barc). May 7 2021;156(9):428–436. doi:10.1016/j.medcli.2020.12.018

53. Kheir M, Saleem F, Wang C, Mann A, Chua J. Higher albumin levels on admission predict better prognosis in patients with confirmed COVID-19. PLoS One. 2021;16(3):e0248358. doi:10.1371/journal.pone.0248358

54. Huang J, Cheng A, Kumar R, et al. Hypoalbuminemia predicts the outcome of COVID-19 independent of age and co-morbidity. J Med Virol. Oct 2020;92(10):2152–2158. doi:10.1002/jmv.26003

55. Rizzo G, Mappa I, Pietrolucci ME, Lu JLA, Makatsarya A, D’Antonio F. Effect of SARS-CoV-2 infection on fetal umbilical vein flow and cardiac function: a prospective study. J Perinat Med. May 25 2022;50(4):398–403. doi:10.1515/jpm-2021-0657

56. Anuk AT, Tanacan A, Yetiskin FDY, et al. Doppler assessment of the fetus in pregnant women recovered from COVID-19. J Obstet Gynaecol Res. May 2021;47(5):1757–1762. doi:10.1111/jog.14726

57. Hessami K, Homayoon N, Hashemi A, Vafaei H, Kasraeian M, Asadi N. COVID-19 and maternal, fetal and neonatal mortality: a systematic review. J Matern Fetal Neonatal Med. Aug 2022;35(15):2936–2941. doi:10.1080/14767058.2020.1806817

